# Structural and Functional Alterations of the Dorsolateral Prefrontal Cortex Across Chronic Pain Cohorts

**DOI:** 10.64898/2026.03.24.26349122

**Authors:** Morihiko Kawate, Saki Takaoka, Yuta Shinohara, Yihuan Wu, Yuki Mashima, Chisato Tanaka, Naho Ihara, Takashige Yamada, Shizuko Kosugi, Kenta Wakaizumi

## Abstract

**Background:** Chronic pain is associated with structural and functional brain alterations, particularly within prefrontal, insular, and cingulate cortices. The dorsolateral prefrontal cortex (DLPFC) shows consistent structural abnormalities across chronic pain conditions, whereas findings on intrinsic functional connectivity (FC) remains inconsistent. Anchoring FC analyses to structural alterations may help identify consistent patterns across chronic pain conditions.

**Methods:** We employed a voxel-based morphometry (VBM)-guided, seed-based resting-state FC approach. Structural and functional MRI data were obtained from patients with chronic neck pain (CNP; n=21) and healthy controls (HC; n=25). Regions showing significant gray matter volume (GMV) differences were used as seeds for whole-brain FC analysis. Associations with pain intensity and pain-related fear were examined. Findings were further evaluated in an independent cohort with chronic primary pain (CPP; n=38).

**Results:** VBM revealed reduced GMV in the left DLPFC in CNP compared with HC, replicated in CPP. Seed-based FC analysis demonstrated reduced connectivity between the left DLPFC and the right hippocampus in CNP, with a similar pattern in CPP. In CNP, GMV in the DLPFC was positively associated with DLPFC-hippocampal connectivity (r = 0.45, 95% CI 0.02 to 0.74, p = 0.043). Reduced DLPFC-hippocampal connectivity was associated with higher activity avoidance (r = -0.50, 95% CI -0.77 to -0.09, p = 0.021), whereas no associations were observed with pain intensity.

**Conclusions:** These findings indicate consistent structural and functional alterations across chronic pain cohorts.

Reduced DLPFC–hippocampal connectivity may reflect altered interactions between prefrontal and hippocampal circuits involved in pain-related cognitive and affective processes.

## Introduction

Structural MRI studies consistently report reduced gray matter volume (GMV) or density in the prefrontal, insular, and cingulate cortices across various chronic pain (CP) conditions. Early voxel-based morphometry (VBM) investigations demonstrated reduced neocortical GMV in chronic back pain^1^, with focal reductions in bilateral dorsolateral prefrontal cortex (DLPFC) and thalamus that scaled with pain duration, suggesting progressive cortical involvement in chronification. Population-based VBM analyses have further corroborated GMV reductions in the ventrolateral and dorsolateral sectors of the prefrontal cortex, dorsal/ventral medial PFC, and anterior insula among individuals with chronic back pain, linking cortical morphology with pain intensity^2^. Additional large cohorts show reduced GMV within anterior/posterior insula, anterior cingulate cortex (ACC), hippocampus, and superior frontal gyrus across multiple chronic pain conditions^3^.

In contrast to structural findings, fewer studies have demonstrated cross-condition convergence in intrinsic functional connectivity (FC) changes across disparate chronic pain types. While multiple investigations implicate large-scale networks such as the default mode network (DMN)^4^, central executive/frontoparietal network (CEN/FPN), and salience network (SN) in chronic pain, meta-analytic syntheses and reviews emphasize heterogeneity in the directionality and loci of FC alterations across conditions and analytic frameworks. For example, disrupted DMN and SN connectivity has been reported in chronic widespread pain and other chronic pain cohorts^5^, yet patterns vary across studies using independent component analysis, seed-to-voxel, graph metrics, or dynamic FC approaches. Recent coordinate-based meta-analyses suggest tripartite network dysfunction involving DMN, CEN, and SN in chronic pain^6^, but simultaneously highlight condition-specific network rearrangements (e.g., musculoskeletal vs. visceral vs. headache pain), underscoring the challenge of defining a unitary functional signature spanning multiple chronic pain etiologies. Systematic evaluations of graph-based connectivity metrics echo this variability, reporting high heterogeneity and low-certainty evidence for global topology changes in chronic pain^7^.

Several methodological and biological factors plausibly account for the weaker consensus in FC relative to structure. First, the FC signal depends on pain state (rest vs. task, spontaneous fluctuations vs. evoked paradigms), analgesic exposure, and comorbid affective symptoms, each of which can modulate network coupling in opposite directions. Second, analytic variation (seed-based vs. ICA vs. graph/dynamic FC), parcellation schemes, and preprocessing pipelines can yield markedly different connectivity estimates and statistical inferences, hindering cross-study harmonization. Third, even within a single network axis, connectivity changes may be disease-specific—for instance, increased FPN–DMN coupling reported in ankylosing spondylitis^8^, which may not generalize to other chronic pain forms. Although the DLPFC, a core region of the FPN, emerges consistently as structurally compromised in chronic pain (including atrophy in chronic back pain and longitudinal reversibility after pain relief)^9^, reports specifically targeting DLPFC-centered FC are comparatively sparse and often context-dependent (e.g., anticorrelation with periaqueductal gray coupling during spontaneous pain fluctuations in trigeminal neuropathy)^10^, leaving it uncertain whether DLPFC connectivity alterations represent a shared functional hallmark across chronic pain conditions.

This study aimed to identify reproducible FC signatures of chronic pain using a VBM-guided, seed-based framework. Given the robustness of structural findings, a principled strategy to uncover shared functional signatures is to anchor FC analyses to cortical loci that show consistent GMV decreases across chronic and then test reproducibility across independent chronic pain cohorts. By using VBM to identify regions of structural alteration and employing those as seeds for whole-brain FC mapping, one can reduce analytic arbitrariness, increase biological plausibility, and directly probe structure–function coupling relevant to pain chronification. To achieve this, we employ a two-step, VBM-guided seed-to-voxel FC framework. First, we identify cortical regions showing significant structural differences between CNP patients and healthy controls using VBM. Next, we use these structurally altered regions as seeds to map whole-brain FC and determine connectivity patterns that differ between groups. To test generalizability, we examine whether the resultant FC signature replicates in an independent cohort of CPP. This approach is aligned with transdiagnostic perspectives showing common prefrontal-insular structural deficits across chronic pain, depression, and anxiety^11, 12^, while recognizing disorder-specific network patterns. We hypothesize that this approach helps bridge structural commonalities with functional diversity and identify consistent structural and functional alterations associated with chronic pain.

## Methods

This study was approved by the Ethics Committee of Keio University School of Medicine (approval number: 20200345) and registered prior to patient enrollment in the University Hospital Medical Information Network Clinical Trials Registry (UMIN-CTR; ID: UMIN000043846; date of registration: 5 April 2021). All participants provided written informed consent prior to enrollment. For the CNP cohort, which consisted of previously collected data, an opt-out procedure was implemented in accordance with institutional guidelines, allowing participants to decline the secondary use of their data.

### Data and Participants

The primary dataset comprised participants with CNP and HC, originally reported in our previous Keio neck pain studies^13, 14^. The CNP cohort included 21 right-handed patients aged between 20 and 64 years, recruited from the outpatient clinic of Keio University Hospital, along with 25 healthy controls matched for age and sex. Patients were eligible if they experienced persistent pain localized to the neck region for at least three months and reported a pain intensity of 4 or higher on the Numerical Rating Scale (NRS; 0–10) at the initial visit. Individuals with psychiatric disorders, structural brain abnormalities, or an inability to maintain a stable head position during MRI scanning due to pain were excluded.

To assess reproducibility and generalizability, we additionally analyzed a separate dataset comprising 38 patients with CPP, whose pain was not restricted to the neck region. These participants were enrolled in an independent study in which pretreatment assessments—including demographic data collection and MRI scanning—were conducted prior to cognitive behavioral therapy. CPP was diagnosed according to ICD-11 criteria, defined as persistent or recurrent pain lasting for at least three months and not fully explained by another chronic secondary pain condition^15^. Similar exclusion criteria were applied to ensure comparability across groups. All MRI scans for CNP, HC, and CPP participants were acquired using the same scanner and imaging protocol, ensuring methodological consistency for structural and functional analyses.

### Measures

Data were obtained through self-administered questionnaires assessing demographic variables (age, sex, body mass index [BMI]), pain characteristics (intensity and duration), and pain-related fear. Participants rated their average pain intensity over the past four weeks using a 100-mm Visual Analog Scale (VAS), where 0 mm indicates “no pain” and 100 mm indicates “worst imaginable pain.” The VAS is widely recognized as a reliable and valid measure for assessing pain intensity in chronic pain populations^16^.

Pain-related fear was assessed only in the CNP cohort using the Japanese version of the 11-item Tampa Scale for Kinesiophobia (TSK-11), which was developed by Matsudaira et al. and has demonstrated excellent psychometric properties (Cronbach’s α = 0.92)^17^. Total scores range from 11 to 44, with higher scores indicating greater kinesiophobia^18^. The TSK-11 comprises two subscales: Activity Avoidance (TSK-AA) and Somatic Focus (TSK-SF). The reliability and construct validity of these subscales have been supported by our previous work (Cronbach’s α = 0.71 and 0.72, respectively)^19^. Notably, the TSK-AA subscale specifically reflects fear of movement and has been shown to correlate with pain intensity and psychological distress in chronic pain populations^20, 21^.

### Generalizability Assessment Using an External Dataset

To evaluate external validity, we examined an independent dataset of participants with CPP involving multiple pain locations (e.g., headache and/or facial pain, neck pain, back pain, limb pain). For the CPP dataset, whole-brain GMV maps and seed-based FC maps were generated using the same procedures as in the primary analysis. Extracted values were compared among HC, CNP, and CPP groups as described in the Statistical Analysis section.

### Sample size calculation

This study represents a secondary analysis of previously collected data, and no a priori sample size calculation was performed. Given its exploratory nature of the analysis, the sample size was determined by data availability, and the findings should be interpreted with caution.

### Statistical Analysis

Demographic and clinical variables were summarized as mean (SD) for normally distributed variables and as median (minimum, maximum) for non-normally distributed variables. Demographic variables were compared between CNP and HC groups using an unpaired t-test for age, and the chi-square test for sex. TSK total and subscale scores were compared between CNP and HC groups using an unpaired t-test.

All structural and functional brain images used in the following analyses were preprocessed using standard pipelines, as detailed in the Supplementary Methods (S1), prior to statistical testing. Voxel-wise group comparisons were conducted using the randomise tool in the FMRIB Software Library (FSL) with 5,000 permutations, adjusting for age and sex. Significant clusters were identified using a fixed t-threshold of 3.0 and a family-wise error (FWE)-corrected p < 0.05. Mean GMV values were extracted from significant clusters for each participant and compared between groups.

For seed-based FC analyses, group-level comparisons were similarly performed using FSL’s randomise tool with 5,000 permutations, including age and sex as covariates. Significant clusters were identified using a fixed t-threshold of 3.0 and FWE-corrected p < 0.05. Mean FC values within significant clusters were extracted for each participant and used in correlation analyses with GMV in regions identified by the VBM results.

Extracted mean GMV and mean FC values from significant clusters were adjusted for age and sex using linear regression prior to correlation analyses with clinical measures—including pain intensity, TSK-AA, and TSK-SF scores— in participants with CNP.

External validity was evaluated in an independent CPP dataset by the same imaging analysis protocol and comparing the three groups using the Kruskal–Wallis test followed by Steel–Dwass tests. All statistical tests were two-tailed, and p < 0.05 was considered statistically significant. Statistical analyses were conducted using JMP® version 17.0 (SAS Institute Inc., Cary, NC, USA) and MATLAB R2018a (MathWorks, Natick, MA, USA). Neuroimaging analyses were performed using FSL, and brain regions identified in the VBM and FC analyses were visualized using BrainNet Viewer^22^.

## Results

Demographic and clinical characteristics of the Keio neck pain study are summarized in Table 1. No significant differences were observed between groups in age or sex. Participants with CNP reported a median pain duration of 54 months and a mean pain intensity of 47.6 on the VAS. Scores on the TSK subscales (TSK-AA and TSK-SF) were significantly higher in the CNP group than in HC group, indicating greater fear of movement.

**Table 1.** Demographic characteristics of participants in the Keio neck pain study.

|  | HC (n = 25) | CNP (n = 21) | *p* value |
| --- | --- | --- | --- |
| Age (year), mean (SD) | 43.9 (10.0) | 45.6 (10.5) | 0.578 |
| Women, n (%) | 17 (68.0) | 15 (71.0) | 0.801 |
| Pain intensity, mean (SD) |  | 47.6 (23.9) |  |
| Pain duration (month), median (min, max) |  | 54 (4, 292) |  |
| TSK, mean (SD) |  |  |  |
| Total | 15.7 (5.4) | 27.4 (4.3) | < 0.001 |
| Activity avoidance | 9.1 (3.5) | 15.3 (3.3) | < 0.001 |
| Somatic focus | 6.6 (2.2) | 12.1 (2.5) | < 0.001 |
| Group comparisons were performed using the unpaired t-test for age and TSK measures, and the chi-square test for sex. HC: healthy controls, CNP: chronic neck pain, TSK: Tampa Scale of Kinesiophobia, SD: standard deviation. |  |  |  |

VBM revealed significantly reduced GMV in the left DLPFC in CNP group compared with HC group (112 voxels; MNI coordinates: x = –46 mm, y = 41 mm, z = 7 mm; t > 3.0, cluster-level FWE-corrected p < 0.001) (Figure 1a). Mean GMV values were 0.46 in HC group and 0.40 in CNP group (Figure 1b).

**Figure 1.**
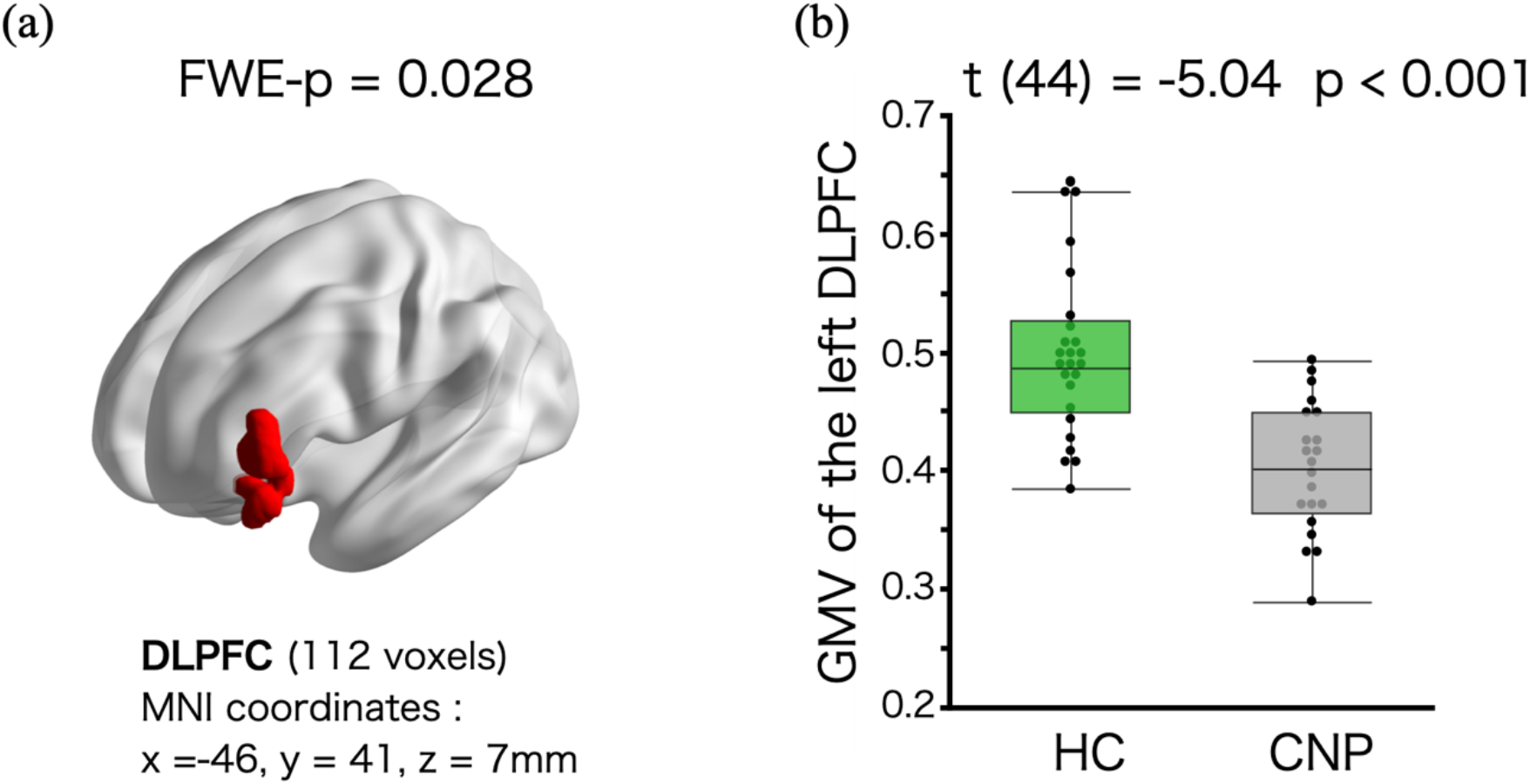
Decreased gray matter volume in participants with chronic neck pain. **(a)** Brain map showing the cluster identified by voxel-based morphometry analysis. **(b)** Group differences in mean gray matter volume of the left dorsolateral prefrontal cortex, assessed using an unpaired t-test. FWE-p: family-wise error corrected p value, HC: healthy controls, CNP: chronic neck pain, DLPFC: dorsolateral prefrontal cortex, MNI: Montreal Neurological Institute, GMV: gray matter volume, p: p value.

Seed-based FC analysis identified a cluster encompassing the right hippocampus that exhibited decreased connectivity in CNP group compared with HC group (697 voxels; MNI coordinates: x = 39 mm, y = –25 mm, z = –22 mm; t > 3.0, cluster-level FWE-corrected p = 0.012) (Figures 2a and 2b). Among CNP participants, GMV in the left DLPFC was positively correlated with FC between the left DLPFC and right hippocampus (r = 0.45, 95% CI 0.02 to 0.74, p = 0.043), indicating that greater structural integrity was associated with stronger FC (Figure 2c).

**Figure 2.**
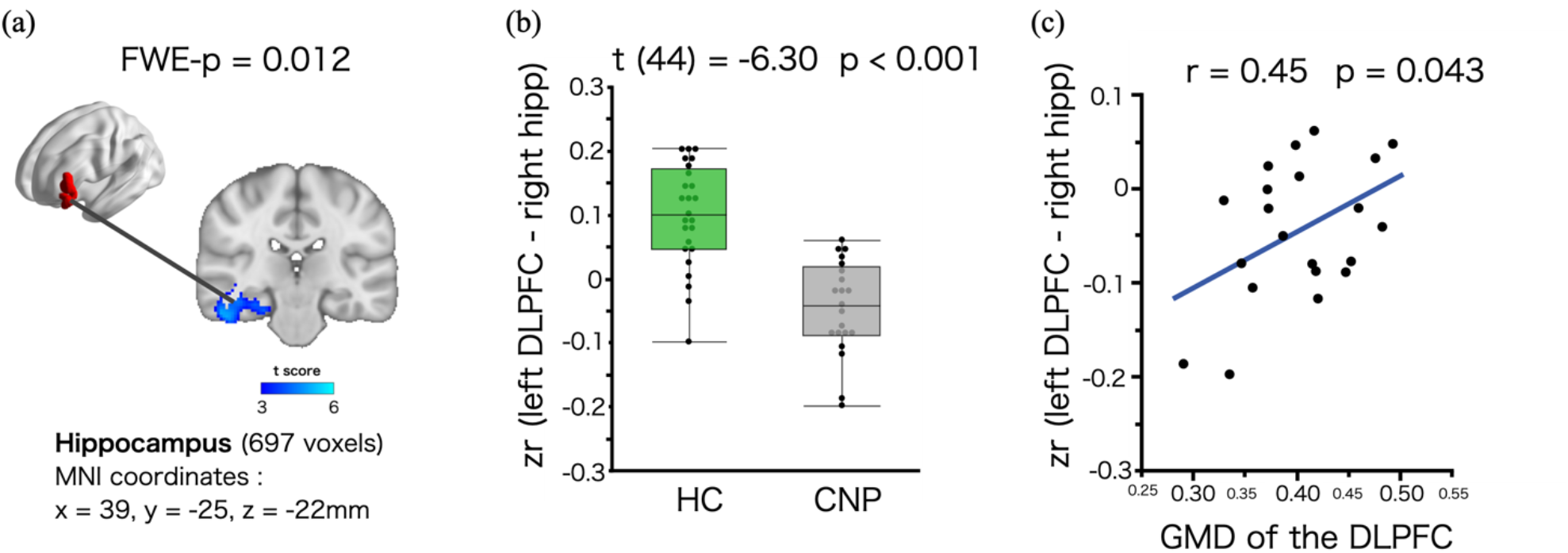
Decreased functional connectivity in participants with chronic neck pain. **(a)** Brain map showing the cluster identified by seed-based FC analysis. **(b)** Group differences in mean FC between the left DLPFC and right hippocampus, assessed using an unpaired t-test. **(c)** Significant association between GMV of the left DLPFC and DLPFC–hippocampal FC. FWE-p: family-wise error corrected p value, FC: functional connectivity, HC: healthy controls, CNP: chronic neck pain, zr: Fisher’s z-transformed correlation coefficient, DLPFC: dorsolateral prefrontal cortex, hipp: hippocampus, GMV: gray matter volume, MNI: Montreal Neurological Institute, p: p value.

After adjusting for age and sex, GMV in the left DLPFC showed no statistically significant associations with pain intensity, TSK-AA, or TSK-SF scores (Figures 3a-3c). However, the correlation between GMV and pain intensity showed a comparable effect size, indicating a similar trend although it did not reach the conventional p < 0.05 threshold. In contrast, FC between the left DLPFC and right hippocampus was negatively correlated with TSK-AA (r = -0.50, 95%CI -0.77 to -0.09, p = 0.021), suggesting reduced connectivity with increased activity avoidance (Figure 3e). Pain intensity and TSK-SF scores did not show significant correlations with this FC measure (Figures 3d and 3f).

**Figure 3.**
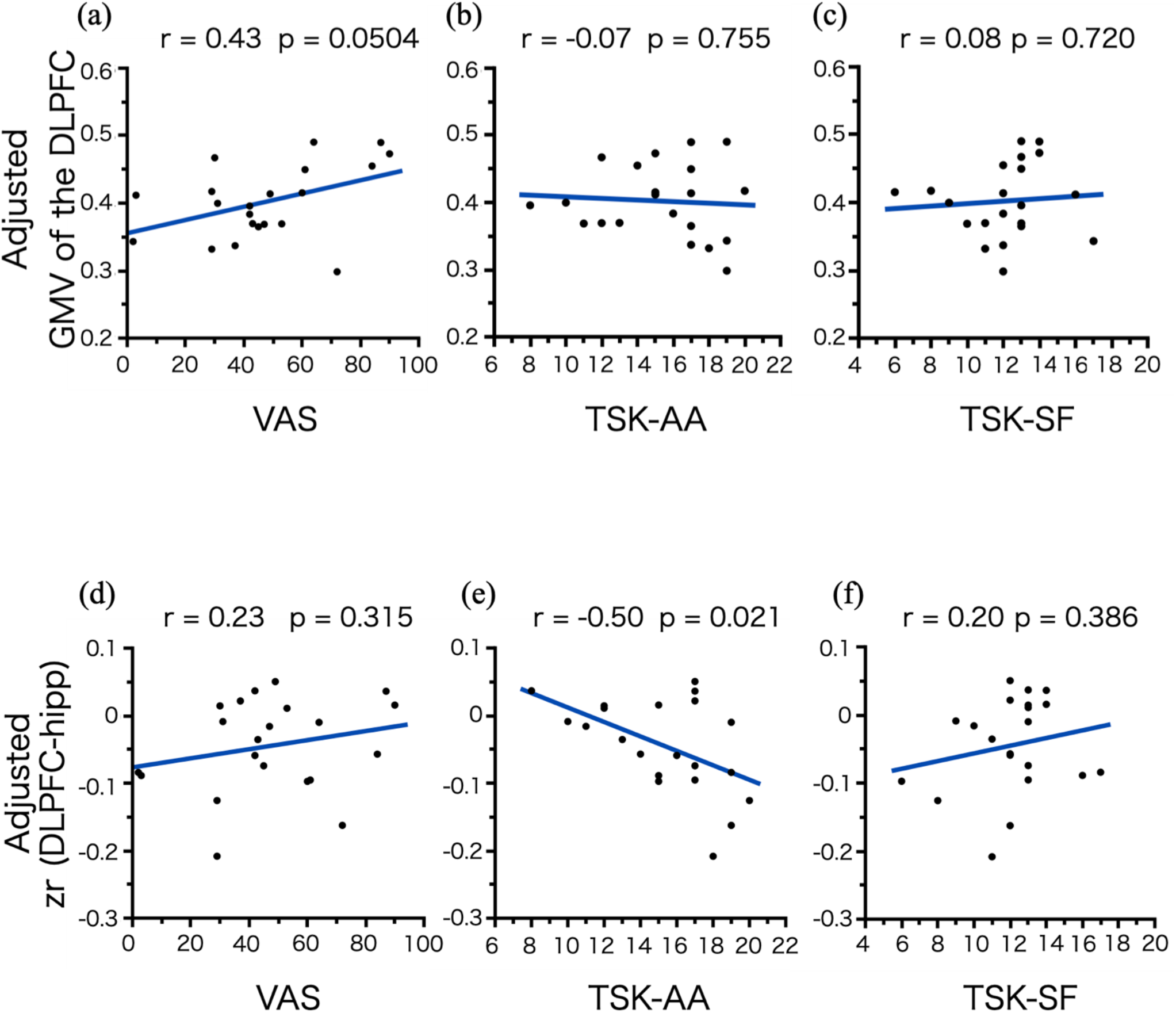
Associations between brain signatures and behavioral measures in participants with chronic neck pain (n = 21). **(a-c)** Scatter plots showing associations of GMV in the DLPFC with pain intensity, TSK-AA, and TSK-SF scores. **(d-f)** Scatter plots showing associations of the DLPFC–hippocampal FC with pain intensity, TSK-AA, and TSK-SF scores. The vertical axes represent adjusted GMV or DLPFC–hippocampal FC values, with age and sex effects removed by linear regression. DLPFC: dorsolateral prefrontal cortex, GMV: gray matter volume, hipp: hippocampus, VAS: visual analogue scale, TSK-AA: Tampa Scale for Kinesiophobia–activity avoidance, TSK-SF: Tampa Scale for Kinesiophobia–somatic focus, zr: Fisher’s z-transformed correlation coefficient.

Demographic and clinical characteristics of the CPP are shown in Table 2. In the CPP group, GMV in the left DLPFC was reduced compared with HC (p = 0.006, Figure 4a). CPP participants also exhibited FC between the left DLPFC, and right hippocampus compared with HC group. There was no statistically significant difference between CNP and CPP groups (p = 0.062, Figure 4b).

**Table 2.** Demographic characteristics of participants with chronic primary pain (n = 38).

|  |  |
| --- | --- |
| Age (years), mean (SD) | 53.1 (13.5) |
| Women, n (%) | 25 (65.8) |
| Pain intensity, mean (SD) | 62.3 (22.0) |
| Pain duration (month), median (min, max) | 64 (5, 659) |
| Pain location, n (%) |  |
| Head and/or face | 14 (36.8) |
| Neck | 10 (26.3) |
| Back | 10 (26.3) |
| Lower limbs | 8 (21.1) |
| Others | 6 (15.8) |
| Upper limbs | 5 (13.2) |
| Normally distributed continuous variables are presented as mean (SD), and non-normally distributed continuous variables are presented as median (min, max). SD: standard deviation. |  |

**Figure 4.**
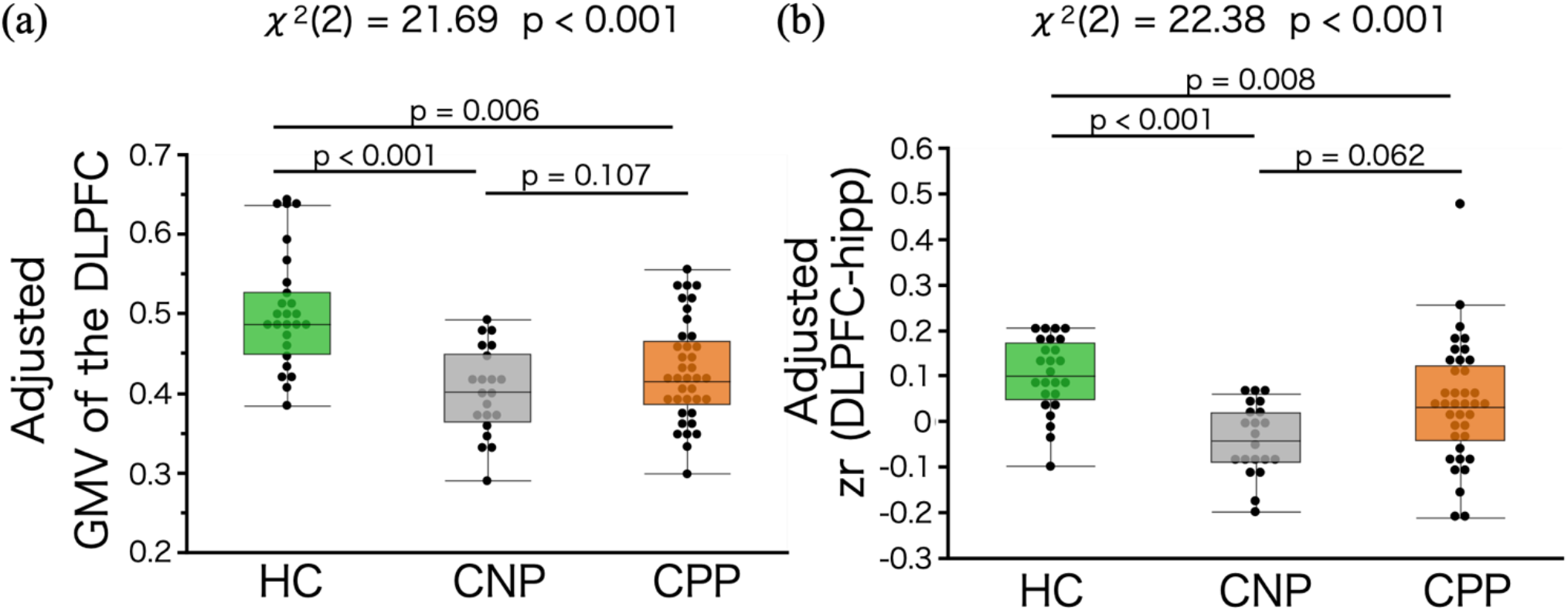
Group comparisons among healthy controls, chronic neck pain, and chronic primary pain. **(a)** Differences in GMV of the left DLPFC, among HC, CNP, and CPP. **(b)** Differences in FC between the left DLPFC and hippocampus among HC, CNP, and CPP. The vertical axes represent adjusted GMV or adjusted DLPFC–hippocampal FC values, with age and sex effects removed by linear regression. Statistical analyses were performed using the Kruskal–Wallis test followed by Steel–Dwass multiple comparison tests. GMV: gray matter volume, DLPFC: dorsolateral prefrontal cortex, FC: functional connectivity, hipp: hippocampus, HC: healthy controls, CNP: chronic neck pain, CPP: chronic primary pain, zr: Fisher’s z-transformed correlation coefficient, p: p value.

## Discussion

The present study investigated whether structural and functional brain alterations associated with CNP, a single well-defined chronic pain condition, are shared across independent chronic pain cohorts. Specifically, we demonstrate convergent structural and functional alterations centered on the DLPFC across two chronic pain phenotypes, CNP and CPP. VBM revealed a reduced GMV in the left DLPFC in CNP compared with HC, and this morphological change was replicated in CPP. Seed-based FC analysis further identified decreased coupling between the left DLPFC and the right hippocampus in CNP, with a similar pattern also observed in CPP. Importantly, higher GMV in the DLPFC was positively associated with stronger FC with the hippocampus, whereas reduced connectivity was correlated with greater activity avoidance measured by the TSK-AA. Notably, no significant association was observed between pain intensity and either GMV in the DLPFC or DLPFC– hippocampal FC. This pattern is consistent with prior chronic pain studies indicating that prefrontal structural and functional alterations are more closely related to maladaptive cognitive and behavioral processes rather than pain intensity^23, 24^.

### Structural Alterations as a Transdiagnostic Feature

Our findings indicate that the DLPFC exhibits consistent structural alterations across distinct chronic pain phenotypes, including CNP and CPP. This observation of similar morphological changes underscores the transdiagnostic nature of DLPFC involvement in chronic pain. Rather than representing condition-specific pathology, these changes likely reflect a common neuroplastic response to prolonged nociceptive input and stress exposure. Functional evidence further supports this interpretation: the DLPFC is engaged during pain suppression and placebo analgesia, consistent with its role in top-down modulation of nociception^25–28^. Structural MRI studies have similarly documented reductions in prefrontal GMV across multiple chronic pain conditions ^1, 29^. Collectively, these findings position the DLPFC as a critical hub affected across chronic pain disorders.

### Fronto-hippocampal Circuit and Behavioral Relevance

The hippocampus is critically involved in fear conditioning, emotional regulation, contextual memory, and nociceptive processing^30^. Structural and FC between the DLPFC and hippocampus have been demonstrated using diffusion tensor imaging and resting-state/task fMRI^31, 32^. Task-based paradigms further show that prefrontal engagement can suppress hippocampal activation, consistent with inhibitory control over unwanted memories^33^. Although previous pain research has primarily focused on hippocampal–medial prefrontal connectivity^34^, our findings extend this literature by demonstrating reduced DLPFC–hippocampal FC in CNP, with replication in CPP. Taken together with decreased GMV in the DLPFC, these results may reflect altered interactions between prefrontal–hippocampal circuits that have been implicated in pain expectancy, contextual appraisal, and stress reactivity.

This decreased connectivity may be related to processes involved in extinction learning and cognitive reappraisal, thereby promoting fear-driven avoidance behaviors. The observed negative association between DLPFC–hippocampal FC and activity avoidance (TSK-AA) underscores the behavioral relevance of this circuit. Together, these findings are consistent with models suggesting altered interactions between prefrontal and hippocampal circuits may be associated with maladaptive pain-related cognition and avoidance.

### Structure–function Coupling and Mechanistic Implications

The positive association between GMV in the DLPFC and its FC with the hippocampus highlights the importance of structure–function coupling in chronic pain. Previous studies suggest that chronic nociceptive input and stress may be associated with dendritic remodeling, synaptic plasticity changes, and alterations in excitatory/inhibitory balance within prefrontal and hippocampal circuits^29, 35^. These changes can impair large-scale networks, including the frontoparietal control network and the salience network, and can bias default mode network dynamics toward pain-related rumination^36^. Structural thinning of the DLPFC may reduce the efficacy of descending pain modulation^37^. In addition, hippocampal dysfunction may sustain maladaptive contextual learning and stress reactivity^30^. Together, these processes are consistent with models proposing that cortical atrophy and network interactions may be related to cognitive control, fear-related learning, and stress-related processes implicated in pain chronification.

### Limitations

This study has several limitations. First, its cross-sectional design precludes any inference about causal relationships among activity avoidance, altered FC, and GMV. Second, the relatively small sample size and single-center recruitment may have introduced selection bias, and psychosocial factors could not be fully controlled. Third, many participants were taking medications such as pregabalin, tramadol, or antidepressants, which may have influenced brain function. Fourth, analyses were based on resting-state fMRI; therefore, network dynamics during pain-related or cognitive tasks were not assessed.

### Future Directions

Future research should include larger and more diverse samples across multiple institutions to improve generalizability. Longitudinal studies are warranted to clarify whether reductions in GMV occur prior to or as a consequence of chronic pain.

## Conclusion

This study demonstrates consistent structural and functional alterations involving the DLPFC across chronic pain cohorts, characterized by structural vulnerability of the DLPFC and reduced functional connectivity with the hippocampus. These alterations were observed in CNP and replicated in CPP, suggesting a potential transdiagnostic involvement of the DLPFC. The association between DLPFC–hippocampal connectivity and activity avoidance highlights the relevance of fronto-limbic circuits in pain-related cognitive and behavioral processes.

## Acknowledgements

The authors used M365 Copilot and ChatGPT 5.2 to assist with English language editing and structural organization during manuscript preparation. All AI-assisted output was reviewed and edited by the authors, who take full responsibility for the accuracy and integrity of the content. No confidential or unpublished patient data were entered into these tools.

## Data Availability Statement

The data presented in this study are available on reasonable request from the corresponding author, subject to approval by the institutional ethics committee due to ethical and privacy restrictions.

## Declarations

### Author Contributions

M.K. was responsible for data analysis, statistical evaluation, interpretation of the results, and drafting of the initial manuscript. S.T. and Y.S. contributed to manuscript editing. Y.W. contributed to editing of the primary draft and assisted with editorial refinement of the text. Y.M. assisted in preparing the initial draft. C.T. critically reviewed the manuscript for important intellectual content. N.I. contributed to data collection and assisted in preparing sections of the preliminary draft. S.K. and T.Y. critically reviewed the manuscript for intellectual content. K.W. conceptualized the study, supervised the project, oversaw data analysis, and provided critical revisions of the manuscript. All authors reviewed and approved the final version of the manuscript.

### Data Availability Statement

The data presented in this study are available on request from the corresponding author due to ethical restrictions.

### Declaration of Generative AI and AI-assisted technologies in the writing process

The authors used M365 Copilot (GPT-5 reasoning) to assist with English language editing and structural organization during manuscript preparation. All AI-assisted output was reviewed and edited by the authors, who take full responsibility for the accuracy and integrity of the content. No confidential or unpublished patient data were entered into the tool.

