## Supplementary methods (S1) for "Structural and Functional Alterations of the Dorsolateral Prefrontal Cortex Across Chronic Pain Cohorts"

Detailed MRI acquisition parameters and preprocessing procedures are described below.

### MRI Data Acquisition

Brain imaging data were acquired using a 3.0-T Signa MRI system (GE Healthcare, Chicago, IL, USA) equipped with an 8-channel phased-array head coil. High-resolution anatomical images were obtained using a three-dimensional T1-weighted sequence with extended dynamic range (brain volume imaging): voxel size = 1 × 1 × 1 mm; inversion time (TI) = 650 ms; flip angle (FA) = 8°; field of view (FOV) = 256 mm.

Resting-state functional images were acquired using a gradient-echo echo-planar imaging sequence in ascending order: slice thickness/gap = 3.2/0.8 mm; FOV = 212 mm; acquisition matrix = 64 × 64; repetition time (TR) = 2500 ms; echo time (TE) = 30 ms; flip angle = 80°; number of slices = 40; total scan duration = 8 minutes. During scanning, participants were instructed to remain awake with their eyes open, avoid excessive movement, and refrain from focusing on any specific thoughts.

**Preprocessing of Structural Images**

Gray matter volume (GMV) was analyzed using voxel-based morphometry (VBM) implemented in FSL-VBM[1], following an optimized VBM protocol[2] with FSL tools[3]. Structural T1-weighted images first underwent brain extraction, followed by segmentation of gray matter. The resulting gray matter maps were then nonlinearly registered to the Montreal Neurological Institute (MNI) standard template[4]. Each participant's native gray matter image was aligned to the study-specific template, and modulation was applied to preserve local volume information after nonlinear registration. Finally, the modulated gray matter maps were smoothed using an isotropic Gaussian kernel ($\sigma$ = 3 mm), corresponding to a full width at half maximum (FWHM) of 7.1 mm.

**Preprocessing of Functional Images**

Resting-state functional MRI (rs-fMRI) data were preprocessed using FSL following a standardized pipeline. Each participant's brain was segmented into gray matter, white matter, and cerebrospinal fluid (CSF). Denoising was performed by regressing out signals from CSF, white matter, global signal, and six head motion parameters. For quality control, framewise displacement (FD), framewise rotation (FR; rotational displacements converted using a 50 mm head radius), DVARS, and framewise signal standard deviation (SD) were computed. Volumes were censored if any of the following criteria were met: FD + FR > 0.5 mm, DVARS z-score ≥ 2.3, or signal SD z-score ≥ 2.3; additionally, one volume immediately before and after each flagged volume was removed.

The retained time series were band-pass filtered using a zero-phase first-order Butterworth filter (0.0075–0.10 Hz; TR = 2.5 s), with padding applied to minimize edge artifacts. After quality control, structural images were nonlinearly registered to the 2 mm isotropic MNI template, and the same transformations were applied to the denoised rs-fMRI data. Normalized functional images were then spatially smoothed using an isotropic Gaussian kernel ($\sigma$ = 3 mm), corresponding to a FWHM of 7.1 mm.
